# The Effect of Pandemic Prevalence on the Reported Efficacy of SARS-CoV-2 Vaccine Candidates: A Systematic Review and Meta-analysis

**DOI:** 10.1101/2021.06.05.21258394

**Authors:** Rajeev Sharma, Abhijith Anand

## Abstract

**Importance:** The efficacy of SARS-CoV-2 vaccine candidates reported in Phase 3 trials varies from ∼45% to ∼95%. It is important to explain the reasons for this heterogeneity.

**Objective:** To test the hypothesis that the efficacy of SARS-CoV-2 vaccine candidates falls with increasing prevalence of the COVID-19 pandemic.

**Data Sources:** *ClinicalTrials.gov*, WHO, McGill and LSHTM trackers of COVID-19 candidate vaccines, peer reviewed publications, and press releases were searched until March 31^st^, 2021.

**Study Selection:** All RCTs reporting efficacy outcomes from Phase 3 trials till March 31^st^, 2021 were included. Of the 11 vaccine candidates that had started their Phase 3 trials by November 1, 2020. Phase 3 efficacy outcomes were available for 8 vaccine candidates. (PROSPERO CRD42021243121).

**Data Extraction and Synthesis:** Both authors independently extracted the data required from identified sources, using PRISMA guidelines. The analysis included all RCTs reported in peer reviewed publications and publicly available sources. A random effects model with restricted maximum likelihood estimator was used to summarize the treatment effects. Cochrane Risk of Bias Assessment Tool was used to assess risk of bias. Certainty of evidence was assessed using the GRADE tool.

**Main Outcomes and Measures:** SARS-CoV-2 infections per protocol in vaccine and placebo groups, risk ratio, prevalence of the COVID-19 infection rate in the populations where the Phase 3 trials were conducted.

**Results:** 8 vaccine candidates had reported efficacy data from a total of 20 independent Phase 3 trials, representing a total of 221,968 subjects, 453 infections across the vaccinated groups and 1,554 infections across the placebo groups. The overall estimate of the risk-ratio is 0.24 (95% CI, 0.17-0.34, p < 0.01), with an I^2^ statistic of 88.73%. The meta-regression analysis with pandemic prevalence as the moderator explains almost half the variance in risk ratios across trials (R^2^=49.06%, p<0.01).

**Conclusion and Relevance:** Pandemic prevalence explains almost half of the between-trial variance in reported efficacies. Efficacy of SARS-CoV-2 vaccine candidates declines as the pandemic prevalence increases.

**Key Points:** *Question:* Does the prevalence of the COVID-19 pandemic explain the heterogeneity in efficacies reported across Phase 3 trials of SARS-CoV-2 vaccine candidates?

*Findings:* Almost 50% of the variance in efficacies reported across Phase 3 trials can be explained by differences in COVID-19 infection rate prevailing across trials. Efficacy of evaluated SARS-CoV-2 vaccine candidates falls significantly with increasing prevalence of the COVID-19 pandemic across trial sites.

*Meaning:* Efficacy of SARS-CoV-2 vaccine candidates needs to be interpreted in conjunction with the prevalence of the COVID-19 pandemic. Adjustment for location-level prevalence analysis would provide better insights into the efficacy results of Phase 3 trials.

## Introduction

The SARS-CoV-2 pandemic has generated an unprecedented effort towards developing vaccines to halt the pandemic. As of March 31, 2021, 8 vaccine candidates had published efficacy data from a total of 20 independent Phase 3 trials (Table 1)^1-5^.

**Table 1:**
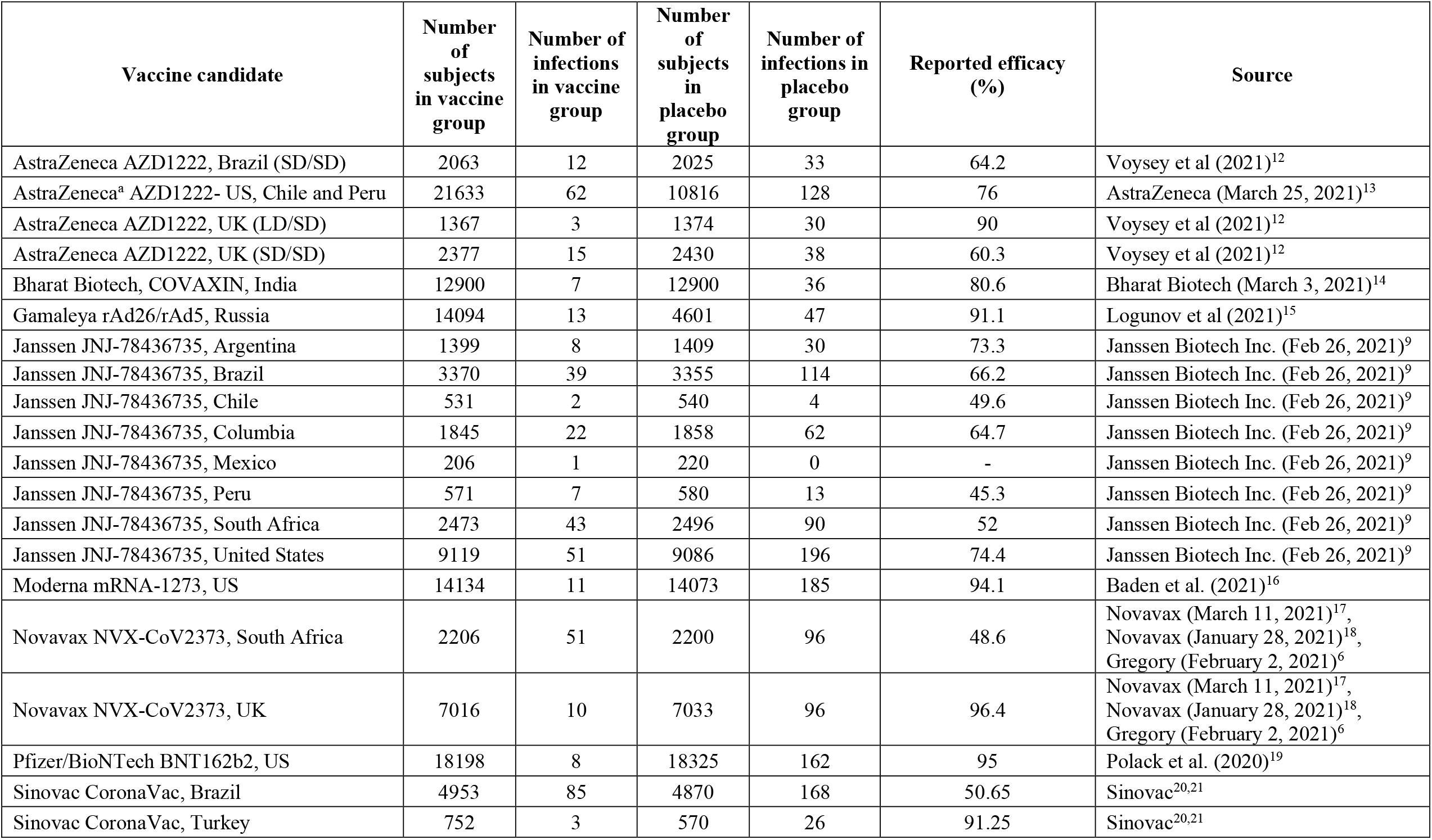
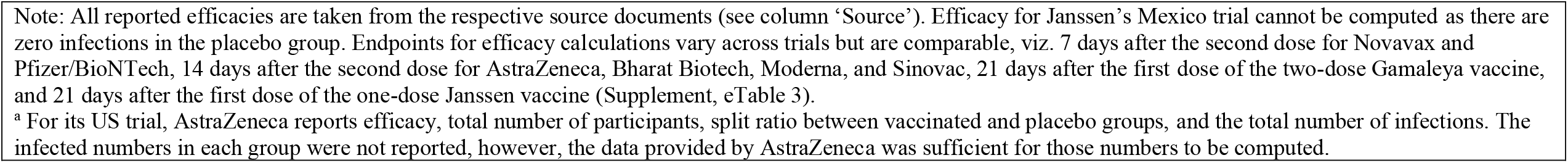
Characteristics of Phase 3 Trials of SARS-CoV-2 Vaccine Candidates Reporting Efficacies.

| Vaccine candidate | Number of subjects in vaccine group | Number of infections in vaccine group | Number of subjects in placebo group | Number of infections in placebo group | Reported efficacy (%) | Source |
| --- | --- | --- | --- | --- | --- | --- |
| AstraZeneca AZD1222, Brazil (SD/SD) | 2063 | 12 | 2025 | 33 | 64.2 | Voysey et al (2021)<sup>12</sup> |
| AstraZeneca<sup>a</sup> AZD1222- US, Chile and Peru | 21633 | 62 | 10816 | 128 | 76 | AstraZeneca (March 25, 2021)<sup>13</sup> |
| AstraZeneca AZD1222, UK (LD/SD) | 1367 | 3 | 1374 | 30 | 90 | Voysey et al (2021)<sup>12</sup> |
| AstraZeneca AZD1222, UK (SD/SD) | 2377 | 15 | 2430 | 38 | 60.3 | Voysey et al (2021)<sup>12</sup> |
| Bharat Biotech, COVAXIN, India | 12900 | 7 | 12900 | 36 | 80.6 | Bharat Biotech (March 3, 2021)<sup>14</sup> |
| Gamaleya rAd26/rAd5, Russia | 14094 | 13 | 4601 | 47 | 91.1 | Logunov et al (2021)<sup>15</sup> |
| Janssen JNJ-78436735, Argentina | 1399 | 8 | 1409 | 30 | 73.3 | Janssen Biotech Inc. (Feb 26, 2021)<sup>9</sup> |
| Janssen JNJ-78436735, Brazil | 3370 | 39 | 3355 | 114 | 66.2 | Janssen Biotech Inc. (Feb 26, 2021)<sup>9</sup> |
| Janssen JNJ-78436735, Chile | 531 | 2 | 540 | 4 | 49.6 | Janssen Biotech Inc. (Feb 26, 2021)<sup>9</sup> |
| Janssen JNJ-78436735, Columbia | 1845 | 22 | 1858 | 62 | 64.7 | Janssen Biotech Inc. (Feb 26, 2021)<sup>9</sup> |
| Janssen JNJ-78436735, Mexico | 206 | 1 | 220 | 0 | - | Janssen Biotech Inc. (Feb 26, 2021)<sup>9</sup> |
| Janssen JNJ-78436735, Peru | 571 | 7 | 580 | 13 | 45.3 | Janssen Biotech Inc. (Feb 26, 2021)<sup>9</sup> |
| Janssen JNJ-78436735, South Africa | 2473 | 43 | 2496 | 90 | 52 | Janssen Biotech Inc. (Feb 26, 2021)<sup>9</sup> |
| Janssen JNJ-78436735, United States | 9119 | 51 | 9086 | 196 | 74.4 | Janssen Biotech Inc. (Feb 26, 2021)<sup>9</sup> |
| Moderna mRNA-1273, US | 14134 | 11 | 14073 | 185 | 94.1 | Baden et al. (2021)<sup>16</sup> |
| Novavax NVX-CoV2373, South Africa | 2206 | 51 | 2200 | 96 | 48.6 | Novavax (March 11, 2021)<sup>17</sup>,<br>Novavax (January 28, 2021)<sup>18</sup>,<br>Gregory (February 2, 2021)<sup>6</sup> |
| Novavax NVX-CoV2373, UK | 7016 | 10 | 7033 | 96 | 96.4 | Novavax (March 11, 2021)<sup>17</sup>,<br>Novavax (January 28, 2021)<sup>18</sup>,<br>Gregory (February 2, 2021)<sup>6</sup> |
| Pfizer/BioNTech BNT162b2, US | 18198 | 8 | 18325 | 162 | 95 | Polack et al. (2020)<sup>19</sup> |
| Sinovac CoronaVac, Brazil | 4953 | 85 | 4870 | 168 | 50.65 | Sinovac<sup>20,21</sup> |
| Sinovac CoronaVac, Turkey | 752 | 3 | 570 | 26 | 91.25 | Sinovac<sup>20,21</sup> |

Note: All reported efficacies are taken from the respective source documents (see column 'Source'). Efficacy for Janssen's Mexico trial cannot be computed as there are zero infections in the placebo group. Endpoints for efficacy calculations vary across trials but are comparable, viz. 7 days after the second dose for Novavax and Pfizer/BioNTech, 14 days after the second dose for AstraZeneca, Bharat Biotech, Moderna, and Sinovac, 21 days after the first dose of the two-dose Gamaleya vaccine, and 21 days after the first dose of the one-dose Janssen vaccine (Supplement, eTable 3).
<sup>a</sup> For its US trial, AstraZeneca reports efficacy, total number of participants, split ratio between vaccinated and placebo groups, and the total number of infections. The infected numbers in each group were not reported, however, the data provided by AstraZeneca was sufficient for those numbers to be computed.

Efficacies reported by the vaccine candidates range from 45% to 95%. Those differences could be due to various sources of heterogeneity between trials^6-8^. At this stage of the vaccine development and emergency use authorization of vaccines based on results of Phase 3 trials, it is important to analyse the available data and account for the effect of potential sources of heterogeneity on vaccine efficacy.

An important source of between-trial heterogeneity that could explain the differences in the efficacies observed across trials is the prevalence of the pandemic in the populations where the trials were conducted^6-9^. The prevalence of the SARS-CoV-2 pandemic has varied across locations and over time. As an example, the prevalence reported in national testing programs has varied from near zero in New Zealand to over 40% in countries such as Mexico and Argentina^10^. Prevalence within countries have also varied over time. For instance, prevalence in the US has varied from a low of 4.0%-5.5% over June 2020 to a high of 10.6%-13.1% over December 2020^10^. Efficacies observed in trials conducted across different countries, and even within a country across different time periods are likely to vary on account of the extent pf prevalence.

Reported efficacies have influenced decisions in critical areas, including those related to public health decisions and individual decisions leading to vaccine hesitancy^11^. To better guide those decisions, we conducted a systematic review and meta-analysis of the efficacy data available from Phase 3 trials. Importantly, we test the hypothesis that as the prevalence of the SARS-CoV-2 pandemic across the trials increases, the observed efficacy will decrease.

## Methods

### Search strategy and selection criteria

This study is a systematic review and meta-analysis of the efficacy data reported in Phase 3 trials of SARS-CoV-2 vaccine candidates. The meta-analysis reports an overall summary estimate of the risk-ratio and conducts a meta-regression to test the effect of between-trial differences in prevalence of the SARS-CoV-2 pandemic on efficacies reported across trial sites.

Vaccine candidates were included in this study if they reported efficacy results from Phase 3 randomised controlled trials. Results of Phase 3 trials reported till March 31, 2021 were included in the analysis.

Phase 3 clinical trials of vaccine candidates are being conducted by the vaccine manufacturers only. We started our search by identifying vaccine candidates that had registered Phase 3 trials, following PRISMA guidelines. Given the high level of interest in the development of SARS-CoV-2 vaccines, such information is widely shared by the manufacturers and tracked by multiple reliable sources^1-3^. In particular, the WHO maintains a tracker database that compiles detailed information on the SARS-CoV-2 vaccine candidate landscape, tracks vaccine candidates in development, and regularly updates progress on registered trials that are underway: “*To ensure the latest information is available, the landscape will be updated twice a week (Tuesday and Friday, 17:00 CET) by searching, gathering and cross-checking data from multiple sources such as the Cochrane vaccine mapping tool, PubMed, ClinicalTrials.gov, WHO ICTRP and from a network of researchers and industry for new candidate vaccines by screening registered trials for clinical information. Where data is missing, … we supplement information gathered from press or public releases*”^1^. Similar information is compiled independently by McGill University’s^2^ and the LSHTM’s Vaccine Centre’s^3^ COVID19 Vaccine Tracker websites. Given the rigorous process followed by those websites, which includes searching of key databases such as *PubMed* and *ClinicalTrials.gov*, we relied on those sources to identify vaccine candidates that had progressed to Phase 3 trials.

The data on vaccine candidates from the three websites was aggregated by RS to compile a master list of vaccine candidates (Supplement, eTable 1). A total of 26 unique vaccine candidates that had progressed to Phase 3 trials were identified. There was 100% concordance between the three tracker websites regarding vaccine candidates currently in Phase 3 trials (Supplement, eTable 1). RS then searched the information available on the three tracker websites, and *ClinicalTrials.gov* to identify the start dates of the Phase 3 trials for each vaccine candidate. Given the time lag between the start of Phase 3 trials and the reporting of efficacy results, trials that had not started by November 1, 2020 were excluded from a further search of results of Phase 3 trials. This left a total of 11 vaccine candidates to be searched for the results of efficacy data from Phase 3 trials. AA independently validated every step of the search, and the master list of vaccine candidates through reference to additional sources (Supplement, eTable 1). No discrepancies were located, and no additional vaccine candidates were located.

RS then searched for original sources where efficacy results of the Phase 3 trials were reported. The progress of vaccine candidates through various trial stages is also of intense public interest and receives unprecedented public scrutiny^1-3^. RS then located links to Phase 3 registered trials from all three tracker websites. Source publications reporting efficacy data for four vaccine candidates were identified on *ClinicalTrials.gov* (AstraZeneca, Gamaleya, Moderna, and Pfizer, see Figure 1). RS then visited the websites of the remaining vaccine manufacturers, which yielded source documents for another four vaccine candidates (Bharat Biotech, Janssen, Novavax, and Sinovac). No efficacy data could be located for the other 3 vaccine candidates, which are from Sinopharm and CanSino, who have chosen not to publicly report the data^22^ (Supplement, eTable 2).

**Figure 1:**
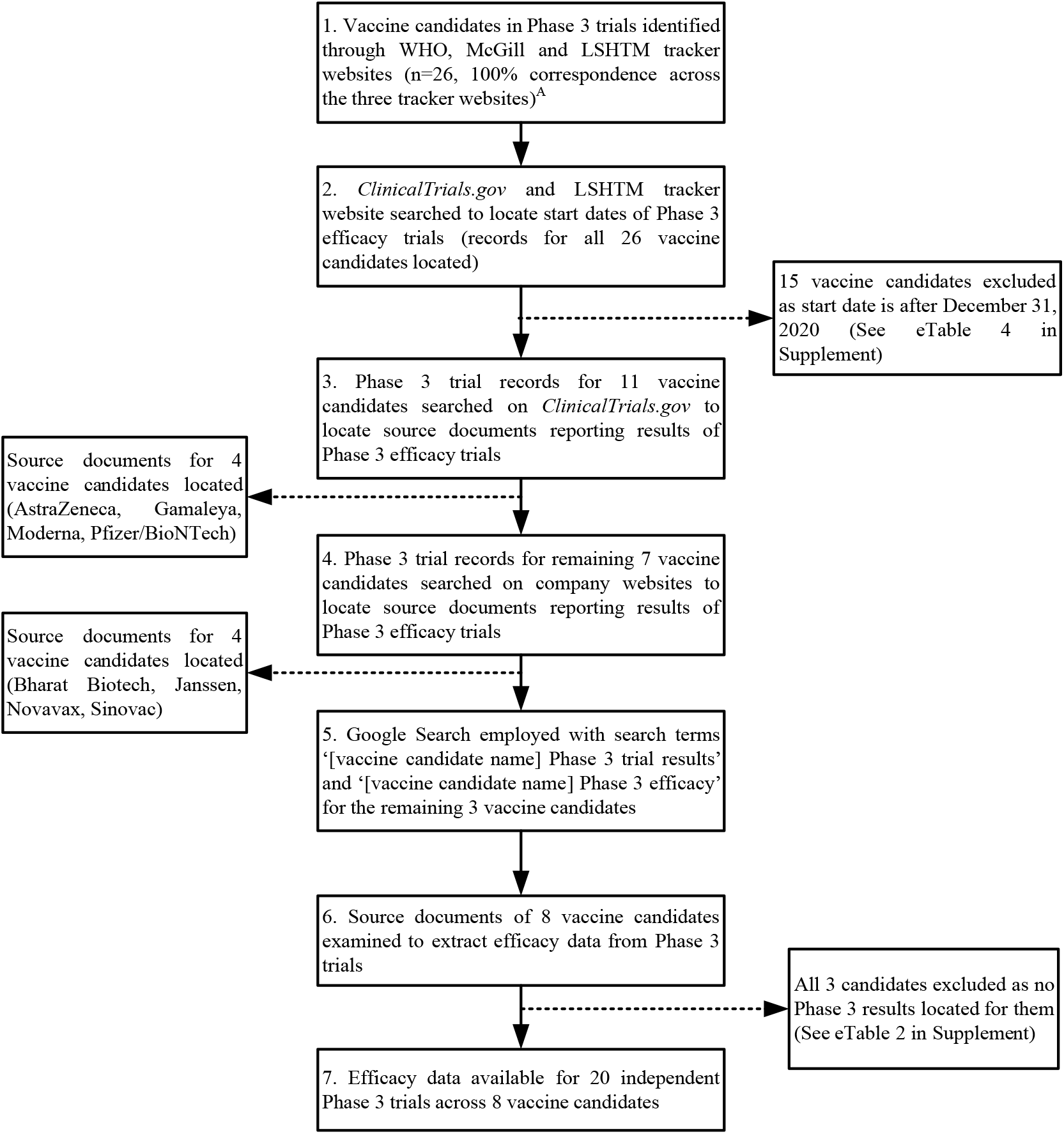
Study Selection.

The search identified 8 vaccine candidates that had reported efficacy data. Table 1 lists the efficacy-related data from 20 independent Phase 3 trials reported by those 8 vaccine candidates, viz. AstraZeneca AZD1222 (4 trials), Bharat Biotech Covaxin, Janssen JNJ-78436735 (8 trials), Moderna mRNA-1273, Novavax NVX-CoV2373 (2 trials), Pfizer/BioNTech BNT162b2, and Sinovac CoronaVac (2 trials). Additional details regarding the trials are reported in Supplement, eTable 3.

### Risk of Bias Assessment

Both authors independently assessed the risk of bias using Cochrane Risk of Bias Assessment Tool. Grading of Recommendations Assessment, Development and Evaluation (GRADE) tool was used to assess the uncertainty of evidence. Disagreements were discussed until consensus was reached.

### Data analysis

Key data needed for the review and meta-analysis was clearly identified in the source documents. Hence, authors of primary studies were not contacted. Quantitative data extracted from the source documents was the number of subjects in the intervention and control arms of the trial, and the number of SARS-CoV-2 infections in the two arms. Summary estimates were extracted from the source documents by both authors working independently. There was 100% agreement between the two authors regarding the number of infections in the vaccine and placebo groups. There were a few minor discrepancies between the two authors on the number of subjects in the vaccine and placebo groups in the trials. These were resolved by mutual discussion and with reference to the source documents. Janssen^9^ reported separate results for eight trials, AstraZeneca^12,13^ reported separate results for four trials, and Novavax^17,18^ and Sinovac^20^ reported results for two trials each. Since the multiple trials reported for a vaccine candidate are independent of each other, they are treated as separate data points in the meta-analysis^23^. Data points were not combined for the analysis. Duplicate data was not encountered in the source documents. Numbers associated with any sub-groups or sub-populations within a trial were not extracted or analysed. The numbers extracted were those reported for the overall trial. No trials whose efficacy results were available were excluded from the analysis.

The primary outcome and the measure of effect analysed was the risk ratio associated with each trial. The risk ratio was computed by the statistical software (STATA16.1) based on the number of infections and participants in the experimental and control arms of the trials. A random-effects model with restricted maximum likelihood estimator was employed in the meta-analysis. Heterogeneity between studies was assessed using the *I*^2^ estimate of heterogeneity^23^.

Pandemic prevalence associated with a trial is estimated from the country-wise data on the SARS-CoV-2 positivity rates reported on Oxford University’s ‘Our World in Data’ portal^10^. The portal reports daily country-wise data for all countries, where available, on the number of people who tested positive for the SARS-CoV-2 infection in the country’s national SARS-CoV-2 testing program^10^. Daily SARS-CoV-2 positive rates corresponding to the duration of each trial were extracted from the portal. The prevalence of the SARS-CoV-2 pandemic associated with each trial was computed as the average of the daily positivity rate prevailing over the duration of the trial (Table 2). Actual start dates for each trial were taken from the trials’ registration data on *ClinicalTrials.gov* and corroborated with the information reported in the peer reviewed documents, company reports and company press releases.

**Table 2:**
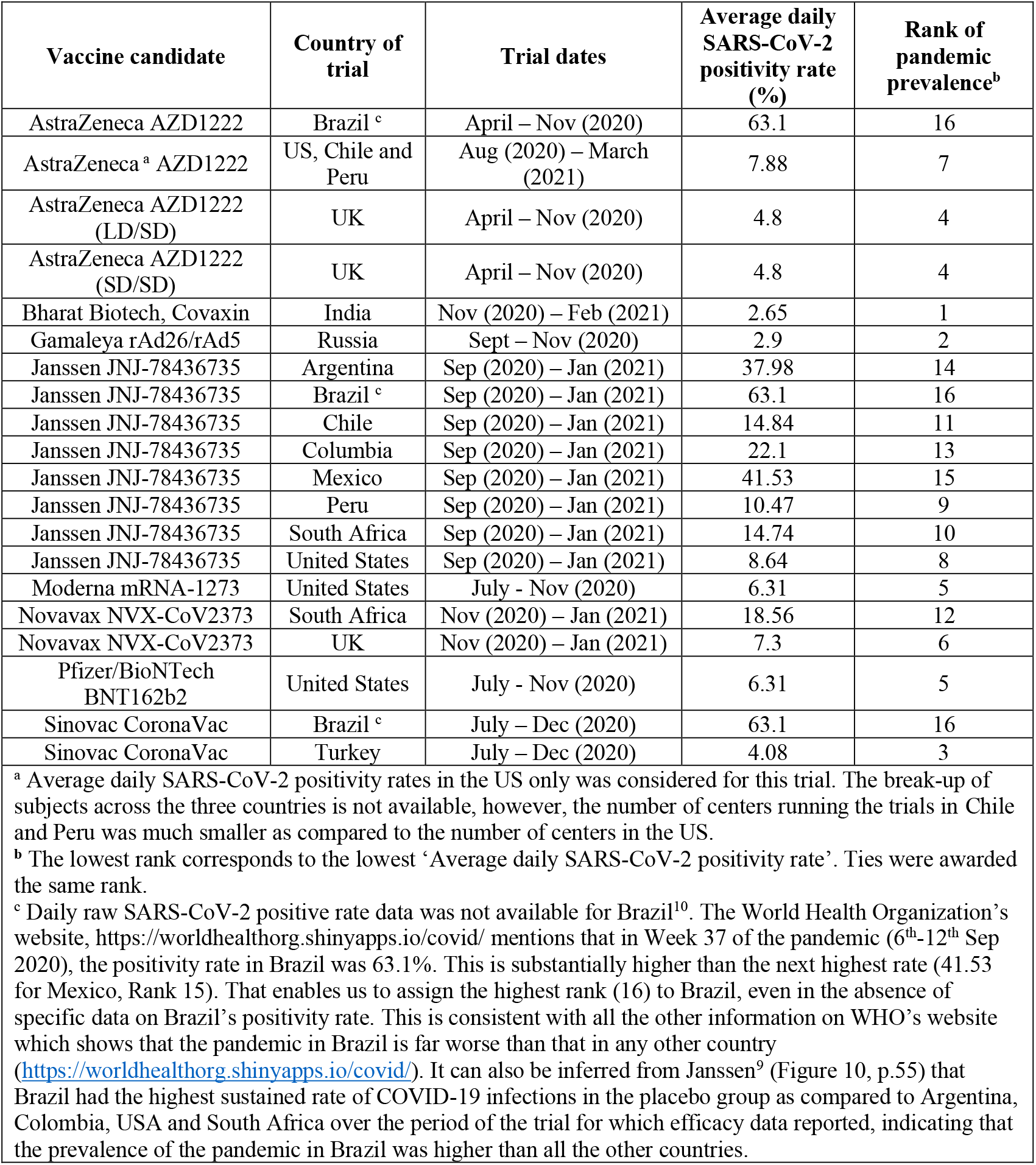
Prevalence of SARS-CoV-2 pandemic associated with trial sites.

| Vaccine candidate | Country of trial | Trial dates | Average daily SARS-CoV-2 positivity rate (%) | Rank of pandemic prevalence<sup>b</sup> |
| --- | --- | --- | --- | --- |
| AstraZeneca AZD1222 | Brazil <sup>c</sup> | April – Nov (2020) | 63.1 | 16 |
| AstraZeneca <sup>a</sup> AZD1222 | US, Chile and Peru | Aug (2020) – March (2021) | 7.88 | 7 |
| AstraZeneca AZD1222 (LD/SD) | UK | April – Nov (2020) | 4.8 | 4 |
| AstraZeneca AZD1222 (SD/SD) | UK | April – Nov (2020) | 4.8 | 4 |
| Bharat Biotech, Covaxin | India | Nov (2020) – Feb (2021) | 2.65 | 1 |
| Gamaleya rAd26/rAd5 | Russia | Sept – Nov (2020) | 2.9 | 2 |
| Janssen JNJ-78436735 | Argentina | Sep (2020) – Jan (2021) | 37.98 | 14 |
| Janssen JNJ-78436735 | Brazil <sup>c</sup> | Sep (2020) – Jan (2021) | 63.1 | 16 |
| Janssen JNJ-78436735 | Chile | Sep (2020) – Jan (2021) | 14.84 | 11 |
| Janssen JNJ-78436735 | Columbia | Sep (2020) – Jan (2021) | 22.1 | 13 |
| Janssen JNJ-78436735 | Mexico | Sep (2020) – Jan (2021) | 41.53 | 15 |
| Janssen JNJ-78436735 | Peru | Sep (2020) – Jan (2021) | 10.47 | 9 |
| Janssen JNJ-78436735 | South Africa | Sep (2020) – Jan (2021) | 14.74 | 10 |
| Janssen JNJ-78436735 | United States | Sep (2020) – Jan (2021) | 8.64 | 8 |
| Moderna mRNA-1273 | United States | July - Nov (2020) | 6.31 | 5 |
| Novavax NVX-CoV2373 | South Africa | Nov (2020) – Jan (2021) | 18.56 | 12 |
| Novavax NVX-CoV2373 | UK | Nov (2020) – Jan (2021) | 7.3 | 6 |
| Pfizer/BioNTech BNT162b2 | United States | July - Nov (2020) | 6.31 | 5 |
| Sinovac CoronaVac | Brazil <sup>c</sup> | July – Dec (2020) | 63.1 | 16 |
| Sinovac CoronaVac | Turkey | July – Dec (2020) | 4.08 | 3 |

<sup>a</sup> Average daily SARS-CoV-2 positivity rates in the US only was considered for this trial. The break-up of subjects across the three countries is not available, however, the number of centers running the trials in Chile and Peru was much smaller as compared to the number of centers in the US.
<sup>b</sup> The lowest rank corresponds to the lowest ‘Average daily SARS-CoV-2 positivity rate’. Ties were awarded the same rank.
<sup>c</sup> Daily raw SARS-CoV-2 positive rate data was not available for Brazil<sup>10</sup>. The World Health Organization’s website, https://worldhealthorg.shinyapps.io/covid/ mentions that in Week 37 of the pandemic (6<sup>th</sup>-12<sup>th</sup> Sep 2020), the positivity rate in Brazil was 63.1%. This is substantially higher than the next highest rate (41.53 for Mexico, Rank 15). That enables us to assign the highest rank (16) to Brazil, even in the absence of specific data on Brazil’s positivity rate. This is consistent with all the other information on WHO’s website which shows that the pandemic in Brazil is far worse than that in any other country (https://worldhealthorg.shinyapps.io/covid/). It can also be inferred from Janssen<sup>9</sup> (Figure 10, p.55) that Brazil had the highest sustained rate of COVID-19 infections in the placebo group as compared to Argentina, Colombia, USA and South Africa over the period of the trial for which efficacy data reported, indicating that the prevalence of the pandemic in Brazil was higher than all the other countries.

The SARS-CoV-2 positivity rates across countries are not directly comparable due to the different protocols and practices across the different national SARS-CoV-2 testing programs^10^. Some of the key differences across countries include different testing regimes, different rates of testing, the different standards of testing, and different reporting practices across countries^10^. To address that limitation, we transformed the raw SARS-CoV-2 positivity rates into ranks for use as a moderator in the meta-regression; while the raw SARS-CoV-2 positivity rates may include a large error component, the ranks are likely to carry a much smaller error component.

The study was registered at PROSPERO (Registration Number CRD42021243121).

## Results

The overall estimate of the risk-ratio (Forest plot reported in Figure 2) is 0.24 (p<0.01; 95% CI: 0.17, 0.34). This equates to an overall efficacy of 76%.

**Figure 2:**
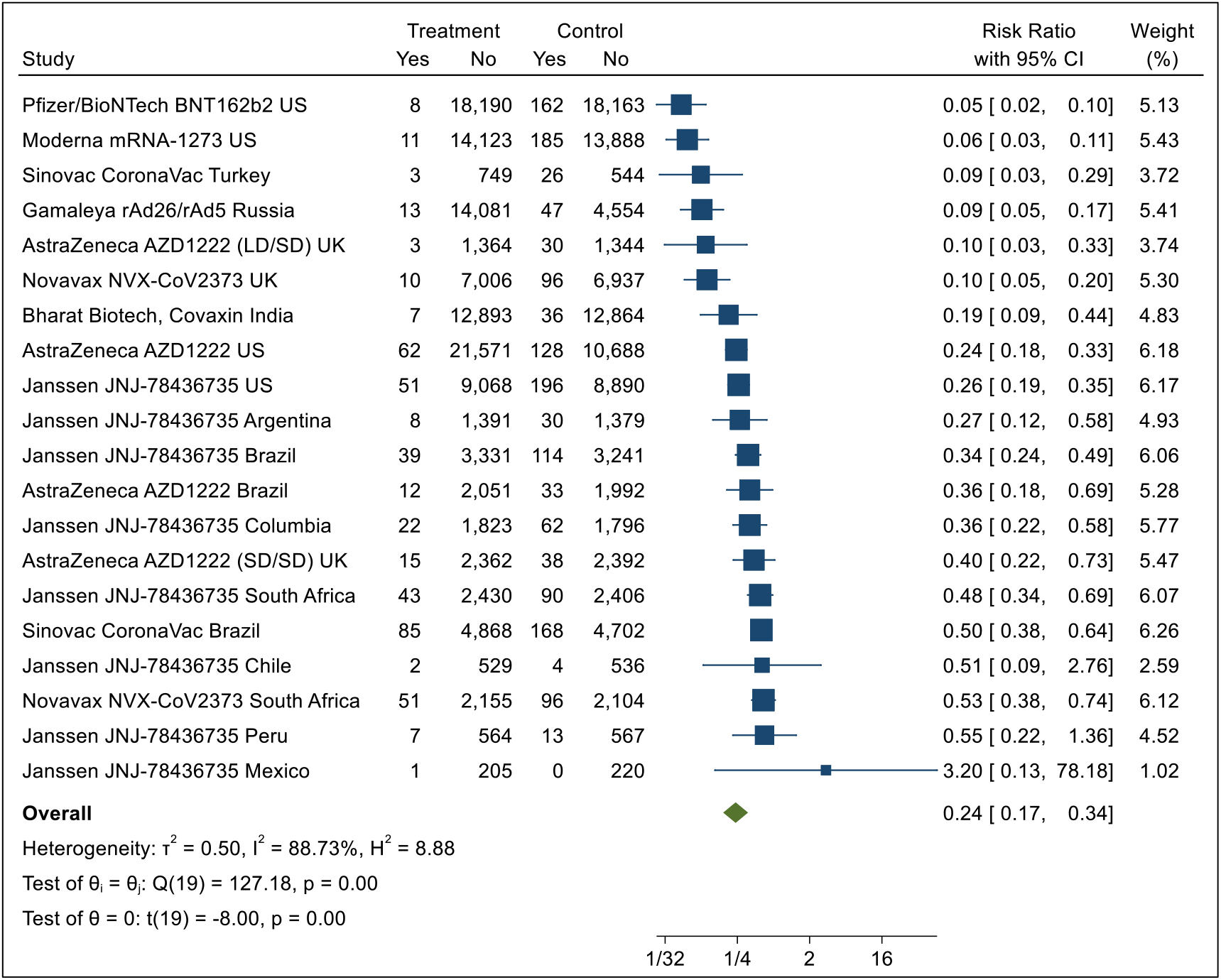
Forest plot.

The I^2^ statistic is 88.73%, suggesting that the extent of heterogeneity is considerable, and that a high proportion of the total variance is due to between-study heterogeneity^24^. This justifies attempts to explain the heterogeneity using between-study differences which, in our case, is prevalence of the pandemic.

Heterogeneity (L’Abbe plot, Supplement eFigure 1) is formally explored via meta-regression with pandemic prevalence as the moderator. A meta-regression of the log risk ratio on pandemic prevalence reports that almost half of the variance in log risk ratios across trials is explained by pandemic prevalence (R^2^=49.06%, p<0.01) (Table 3). The inverse relationship between pandemic prevalence and observed vaccine efficacy hypothesised in this study is supported.

**Table 3:** Results of Meta-regression.

|  | Coefficient | Standard Error | t | P>t | [95%Conf. Interval] |  |
| --- | --- | --- | --- | --- | --- | --- |
| Pandemic prevalence | 0.105 | 0.028 | 3.71 | 0.002 | 0.046 | 0.165 |
| Constant | -2.371 | 0.291 | -8.15 | 0.000 | -2.982 | -1.760 |

Note: Dependent Variable = Log Risk Ratio, Residual heterogeneity $I^2 = 79.3\%$ , $H^2 = 4.83$ , R-squared= 49.06%, Test of residual homogeneity: $Q_{res} = \chi^2(18) = 68.43$ , $Prob>Q_{res} = 0.0000$

The funnel plot (Supplement, eFigure 2) shows no visually obvious small-study effect on the findings. Results of the Egger test report no small-study effects on the findings (beta1 = 0.28, p = 0.73), supporting the visual interpretation (Supplement, eTable 4). Results of nonparametric trim-and-fill analysis of publication bias reports no publication bias due to missing studies; estimated number of imputed studies = 0 (Supplement, eTable 5).

Sensitivity analyses were conducted to analyse if the result of the meta-regression is sensitive to the inclusion of multiple trials for the Janssen vaccine (8 trials) or the AstraZeneca vaccine (4 trials). Two meta-regressions were run, one with an additional dummy variable for the Janssen vaccine, and the other with an additional dummy variable for the AstraZeneca vaccine. In both cases, the regression coefficient for pandemic prevalence was significant, while the coefficients for the dummy variable (Janssen/AstraZeneca) were non-significant (Supplement, eTables 6 and 7).

Another sensitivity analysis was run excluding the data points for Brazil. This was done as the daily SARS-CoV-2 positivity rate for Brazil was not available as its rank of pandemic prevalence was estimated indirectly (see Table 2, Note c). Excluding those three trials does not change the result of the meta-regression; the effect of pandemic prevalence remains significant (R^2^ = 44.56%, p < 0.01) (Supplement, eTable 8). Risk of bias was assessed as low using the Cochrane tool, while certainty of evidence was assessed as high using GRADE (Supplement, eTables 9,10).

## Discussion

The meta-analysis (Forest plot, Figure 2) finds that the overall risk ratio across all 8 vaccines, reporting a total of 20 trials, is 0.24 (p<0.01; 95% CI: 0.17, 0.34), corresponding to an overall efficacy of 76%. The overall efficacy, 76%, is substantially higher than the threshold of 30% often considered as acceptable in Phase 3 trials^16,19^. Overall, the vaccines demonstrate high efficacy.

The results support our hypothesis that increasing pandemic prevalence leads to lower efficacy of SARS-CoV-2 vaccine candidates (Table 3, R^2^=49.06%, p<0.05). Support for the hypothesis is robust, and not sensitive to potential sources of validity threats. The inclusion of multiple trials for the Janssen and AstraZeneca vaccine candidates does not affect the results of the meta-regression reported in Table 3. The finding reported in Table 3 is also robust against the inclusion of trials conducted in Brazil.

The key implication of this study is that a substantial proportion of the differences in efficacy observed across trials can be attributed to the prevalence of the SARS-CoV-2 pandemic across trial sites. This one source of heterogeneity explains almost 50% of the variance in efficacies observed across trials. There are a number of other sources of heterogeneity that occur across Phase 3 trials of the SARS-CoV-2 vaccine candidates too, including, but not limited to doses, interval between doses and the definition of primary endpoints^7,8^. Future research could investigate the effect of other between-trial differences that could explain additional heterogeneity in observed efficacy. Table 3 reports that the I^2^ statistic for residual heterogeneity is 79.3%. This suggests that it is quite likely that the residual heterogeneity could be explained by those additional between-trial differences.

The meta-regression analysis (Table 3) reports that vaccine efficacy falls as pandemic prevalence increases. A key implication of this finding is that the proper interpretation of efficacies reported by vaccine candidates needs to account for the effect of pandemic prevalence.

The finding that pandemic prevalence affects vaccine efficacy suggests two implications for the conduct of Phase 3 trials of SARS-CoV-2 vaccine candidates. First, when efficacy results of Phase 3 trials are reported, they should be reported in conjunction with the level of pandemic prevalence associated with the trial sites. Given the findings of this meta-analysis, a meaningful interpretation of observed efficacy in Phase 3 trials can only be done in conjunction with the associated data on pandemic prevalence.

Second, it suggests that protocols for the design and conduct of Phase 3 trials need to take pandemic prevalence into account. The design of trials should include randomization of trial subjects across locations that vary in the prevalence of the pandemic. Further, for the duration of the Phase 3 trials, manufacturers should monitor pandemic prevalence in all locations where the trials are conducted.

A key strength of this study is that it summarizes the entire current publicly available global evidence on the efficacy of SARS-CoV-2 vaccine candidates. It is also a strength of this study that it can explain almost half the variance in between-trial efficacies with one moderator variable only, pandemic prevalence. That data on pandemic prevalence is obtained from sources independent of the trials, is another strength of this meta-analysis.

### Limitations

The overall sample is small, 20 trials across 8 vaccine candidates. Efficacy data on three other vaccine candidates that have completed Phase 3 trials and are currently approved for use is not publicly available and could not be included in this meta-analysis. All trials are multi-location, and a few are multi-country too, e.g., Pfizer/BioNTech’s trial conducted in the US, Chile and Peru (see Table 2, Note a). In the absence of data on the exact locations of those trials and the number of subjects in the trial at each location, we have used country-level data to rank each trial on pandemic prevalence. This assumption necessarily introduces an element of error in estimating pandemic prevalence. To some extent, that effect is mitigated in our meta-regression by employing rank of pandemic prevalence as the moderator variable. Future Phase 3 trials could be required to report a location-level analysis to provide better insights into the efficacy results of Phase 3 trials.

## Conclusion

Pandemic prevalence has a significant negative effect on efficacies observed across Phase 3 trials of SARS-CoV-2 vaccine candidates; trials conducted in locations with low pandemic prevalence reported higher efficacies as compared to trials conducted in high pandemic prevalence locations.

## Data Availability

All data used in the manuscript is included in the manuscript and supplementary material.

## Author Contributors

RS conceived and conceptualised the research. RS and AA participated in the data collection, validation and analysis. RS and AA jointly wrote the Method and Results sections. RS wrote the other sections, with contributions from AA. Both authors reviewed and edited the manuscript. Both authors have read and agreed to the published version of the manuscript. All authors had full access to all the data in the study and had final responsibility for the decision to submit for publication.

## Data sharing

All datasets generated and analysed are available in the Article and the Supplement.

## Supplemental Content

**eTable 1:**
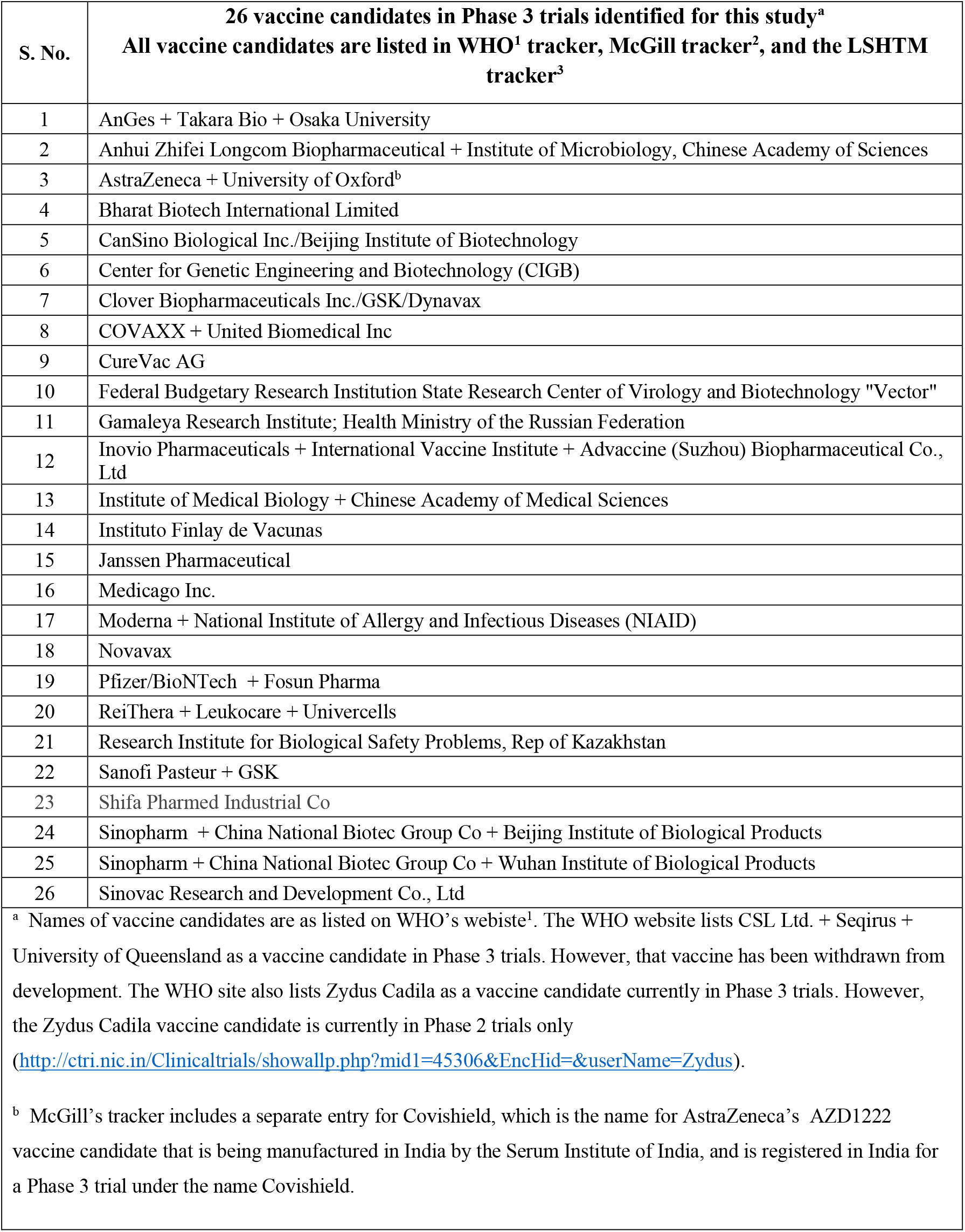
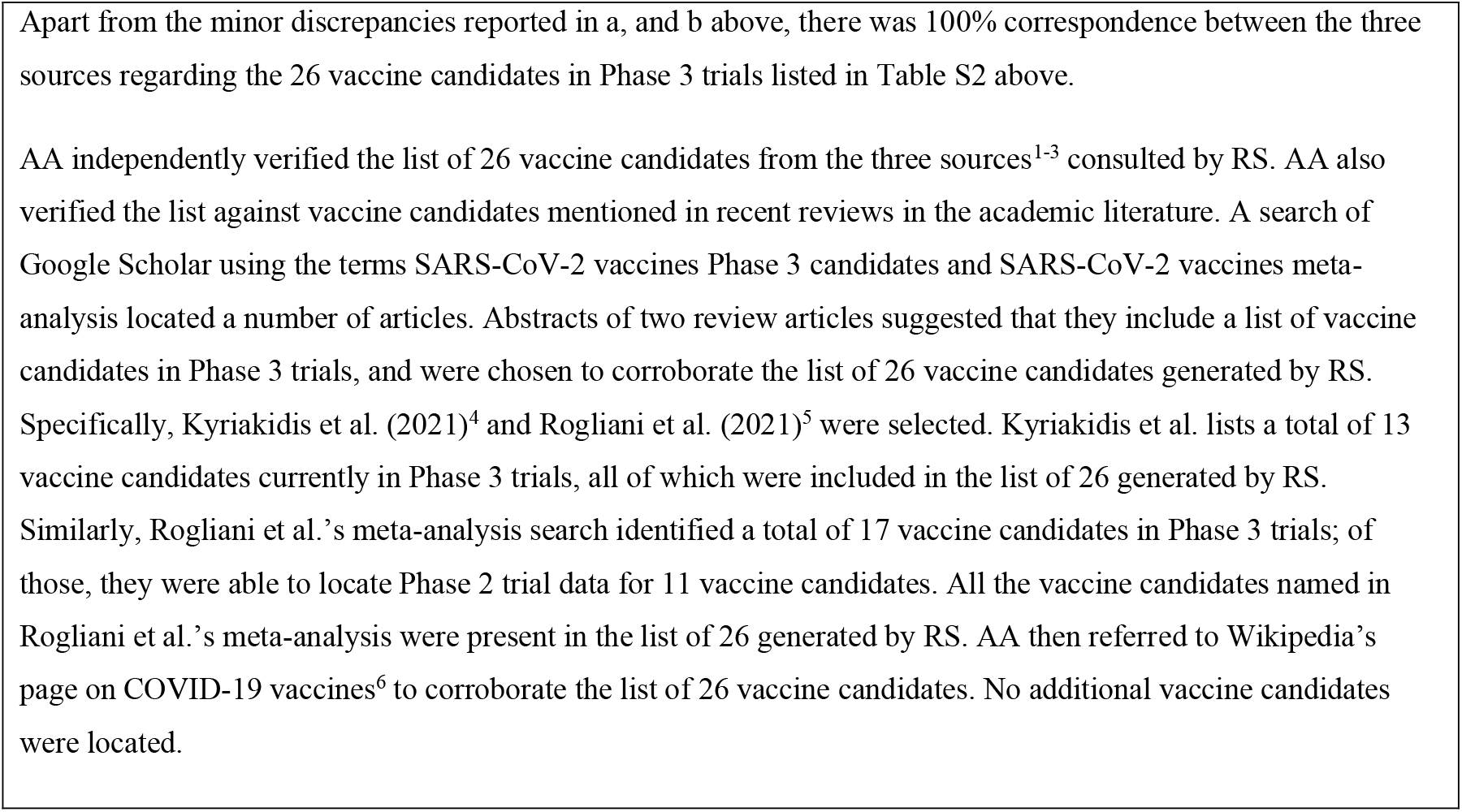
Complete list of 26 vaccine candidates located in Phase 3 trials identified for this study.

**eTable 2:** Search for results of Phase 3 trials for the 26 vaccine candidates.

| **Vaccine Developer** | **Results available on NCT register** | **Results available on company website** | **Company website searched (Outcome of search)** | **Result on Google search** |
| --- | --- | --- | --- | --- |
| AstraZeneca + University of Oxford | Y | Y | “76% vaccine efficacy against symptomatic COVID-19”<br>(https://www.astrazeneca.com/media-centre/press-releases/2021/azd1222-us-phase-iii-primary-analysis-confirms-safety-and-efficacy.html) | “76% vaccine efficacy against symptomatic COVID-19”<br>(https://www.bloomberg.com/press-releases/2021-03-25/azd1222-us-phase-iii-primary-analysis-confirms-safety-and-efficacy) |
| Bharat Biotech International Limited | N | Y | “Bharat Biotech Announces Phase 3 Results of COVAXIN®: India's First COVID-19 Vaccine Demonstrates Interim Clinical Efficacy of 81%.”<br>(https://bharatbiotech.com/press_releases.html) | “India's Covaxin, the homegrown government-backed vaccine, has an efficacy rate of 81%, preliminary data from its phase 3 trial shows.”<br>(https://www.bbc.com/news/world-asia-india-55748124) |
| Gamaleya Research Institute; Health Ministry of the Russian Federation | Y | N | No Information<br>(https://gamaleya.org/en/research/vaktsina-protiv-covid-19/) | “The team found that starting 21 days after the first dose of the vaccine, the vaccine efficacy is 91.6 %”<br>(https://www.chemistryviews.org/details/news/11287590/Preliminary_Phase_3_Trial_Results_of_the_Russian_COVID-19_Vaccine_Sputnik_V.html)<br>“The first interim data analysis of the Sputnik V vaccine against COVID-19 phase III clinical trials in the Russian Federation demonstrated 92% efficacy”<br>(https://www.sciencemag.org/sites/default/files/The%20first%20interim%20data%20analysis%20of%20the%20Sputnik%20V%20vaccine%20against%20COVID-19%20phase%20III%20clinical%20trials%20in%20the%20Russian%20Federation%20demonstrated%2092%25%20efficacy%20.pdf) |
| Janssen Pharmaceutical | Y | Y | Covid19 status webpage (https://www.janssen.com/covid19) (https://www.janssen.com/johnson-johnson-announces-single-shot-janssen-covid-19-vaccine-candidate-met-primary-endpoints) | "...the vaccine was found to be 72 percent effective at preventing what the company defined as moderate to severe Covid-19" (https://www.nbcnews.com/health/health-news/how-effective-johnson-johnson-vaccine-what-know-n1259652) |
| Moderna + National Institute of Allergy and Infectious Diseases (NIAID) | Y | Y | (...Phase 3 Trial of the Moderna COVID-19 Vaccine...) (https://investors.modernatx.com/news-releases/news-release-details/moderna-announces-publication-results-pivotal-phase-3-trial) | "95% efficacy for COVID vaccines doesn't mean there's a 5% infection risk" (https://www.sacbee.com/news/coronavirus/article250559589.html) |
| Novavax | Y | Y | "Novavax COVID-19 Vaccine Demonstrates 89.3% Efficacy..." (https://ir.novavax.com/news-releases/news-release-details/novavax-covid-19-vaccine-demonstrates-893-efficacy-uk-phase-3) | "Novavax's (NVAX) COVID-19 Vaccine Shows Final Efficacy of 96.4%" (https://finance.yahoo.com/news/novavacs-nvax-covid-19-vaccine-130301820.html) |
| Pfizer/BioNTech + Fosun Pharma | Y | Y | "Pfizer and Biontech announce publication of results from landmark phase 3 trial..." (https://www.pfizer.com/news/press-release/press-release-detail/pfizer-and-biontech-announce-publication-results-landmark) | "Pfizer and BioNTech have reported that updated topline results from Phase III study of their Covid-19 vaccine, BNT162b2, demonstrated an efficacy of 91.3% against the disease..." (https://www.pharmaceutical-technology.com/news/pfizer-biontech-vaccine-efficacy/) |
| Sinovac Research and Development Co., Ltd | Y | Y | "Summary of Clinical Trail Data of Sinovac's COVID-19 Vaccine" (http://www.sinovac.com/?optionid=754&auto_id=927) | "A coronavirus vaccine developed by China's Sinovac has been found to be 50.4% effective..." (https://www.bbc.com/news/world-latin-america-55642648) |
| Sinopharm + China National Biotech Group Co + Wuhan Institute of Biological Products | N | N | No information (http://www.sinopharm.com/1156.html) | "No detailed efficacy data of Sinopharm's COVID-19 vaccine has been publicly released" (https://www.reuters.com/article/us-health-coronavirus-who-china-vaccines-idUSKBN2BN1K8) |
| CanSino Biological Inc./Beijing Institute of Biotechnology | N | N | No information<br>(http://www.cansinotech.com/) | "Cansino has not published its phase III trial data in a peer-reviewed journal, medicine's gold standard of vetting. In fact, all four Chinese vaccines on the market have failed to take this step"<br>(https://fortune.com/2021/03/15/covid-vaccine-cansino-china-data/) |
| Sinopharm + China National Biotech Group Co + Beijing Institute of Biological Products | N | N | No information<br>(http://www.sinopharm.com/1156.html) | "No detailed efficacy data of Sinopharm's COVID-19 vaccine has been publicly released" (April 1, 2021)<br>(https://www.reuters.com/article/us-health-coronavirus-who-china-vaccines-idUSKBN2BN1K8) |
| Federal Budgetary Research Institution State Research Center of Virology and Biotechnology "Vector" | N | N | No information,<br>(http://www.vector.nsc.ru/) | "So far, however, researchers have released no corroborating data to the public — not in patents filed for the vaccine or in any other sources. Additionally, their claims about EpiVacCorona's efficacy contradict the findings of an independent laboratory that analyzed blood samples from clinical-trial volunteers and found no neutralizing antibodies." (https://meduza.io/en/feature/2021/03/23/epivaccorona-s-race-to-the-finish-line)<br>"The published research concerns EpiVacCorona's combined Phase I and II trials (involving 14 and 86 volunteers, respectively), meaning that the data reflects only the drug's short-term safety and side effects, as well as its ability to provoke an immune response in patients. These experiments weren't designed to test the vaccine's effectiveness, which can't be assessed until the end of Phase III, which involves 3,000 participants."<br>(https://meduza.io/en/feature/2021/03/27/sputnNik-v-s-ugly-cousin) |
| Medicago Inc. | N | N | "pleased to announce the start of Phase 3 clinical testing"<br>https://www.medicago.com/en/medi-a-room/medicago-and-gsk-start-phase-3-trial-of-adjuvanted-covid-19-vaccine-candidate/) | No Phase 3 results located |
| AnGes + Takara Bio + Osaka University | N | N | "Completion of inoculation in Phase 2/3 trial"<br>(https://www.anges.co.jp/ir/) | No Phase 3 results located |
| Inovio Pharmaceuticals + International Vaccine Institute + Advaccine (Suzhou) Biopharmaceutical Co., Ltd | N | N | "In first quarter 2021, INOVIO completed enrollment of 400 subjects in the Phase 2 segment" (http://ir.inovio.com/news-releases/news-releases-details/2021/INOVIO-Reports-Fourth-Quarter-2020-and-Year-End-Financial-Results/default.aspx) | "completed enrollment of 400 participants in its Phase II segment" (https://www.biospace.com/article/inovio-expects-late-stage-covid-19-vaccine-trial-data-by-second-quarter/) |
| Sanofi Pasteur + GSK | N | N | "“We are pleased to be starting this new Phase 2 study ... We look forward to further progressing this vaccine candidate to Phase 3 in Q2 2021” (https://www.sanofi.com/en/media-room/press-releases/2021/2021-02-22-11-40-00#) | No Phase 3 results located |
| CureVac AG | N | N | "trial will start with an initial Phase 2b part, which is expected to seamlessly merge into a Phase 3 efficacy trial" (https://www.curevac.com/en/2020/12/14/curevac-commences-global-pivotal-phase-2b-3-trial-for-covid-19-vaccine-candidate-cvncov/) | "Data from both clinical trials is expected in the second quarter of 2021" (https://www.pmlive.com/pharma_news/curevac_expands_covid-19_vaccine_trial_protocol_to_include_variant_specification_1365731) |
| Anhui Zhifei Longcom Biopharmaceutical + Institute of Microbiology, Chinese Academy of Sciences | N | N | No website located for Anhui | "There is no publicly available information in peer-reviewed scientific journals about the clinical trial data showing efficacy or safety." (https://www.thehindu.com/news/international/china-approves-fourth-covid-19-vaccine-for-emergency-use/article34080651.ece) |
| Research Institute for Biological Safety Problems, Rep of Kazakhstan | N | N | No website located | No Phase 3 results located |
| Institute of Medical Biology + Chinese Academy of Medical Sciences | - | - | - | Not searched. Trial start date later than 31 December 2020. |
| COVAXX +<br>United Biomedical<br>Inc | - | - | - | Not searched. Trial start date later than 31 December 2020. |
| Clover<br>Biopharmaceutical<br>s<br>Inc./GSK/Dynavax | - | - | - | Not searched. Trial start date later than 31 December 2020. |
| Instituto Finlay de<br>Vacunas | - | - | - | Not searched. Trial start date later than 31 December 2020. |
| Shifa Pharmed<br>Industrial Co | - | - | - | Not searched. Trial start date later than 31 December 2020. |
| ReiThera +<br>Leukocare +<br>Univercells | - | - | - | Not searched. Trial start date later than 31 December 2020. |
| Center for Genetic<br>Engineering and<br>Biotechnology<br>(CIGB) | - | - | - | Not searched. Trial start date later than 31 December 2020. |

**eTable 3:**
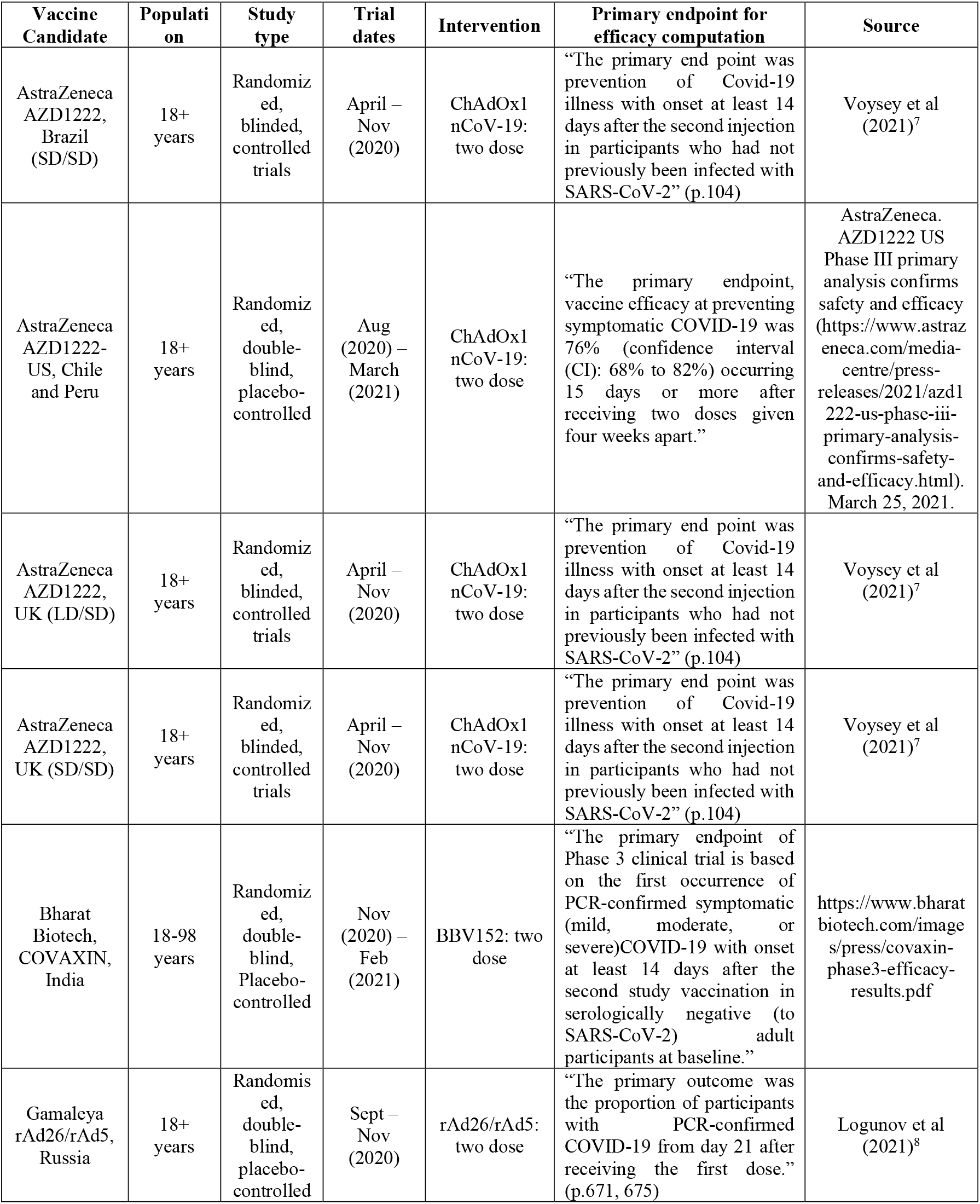
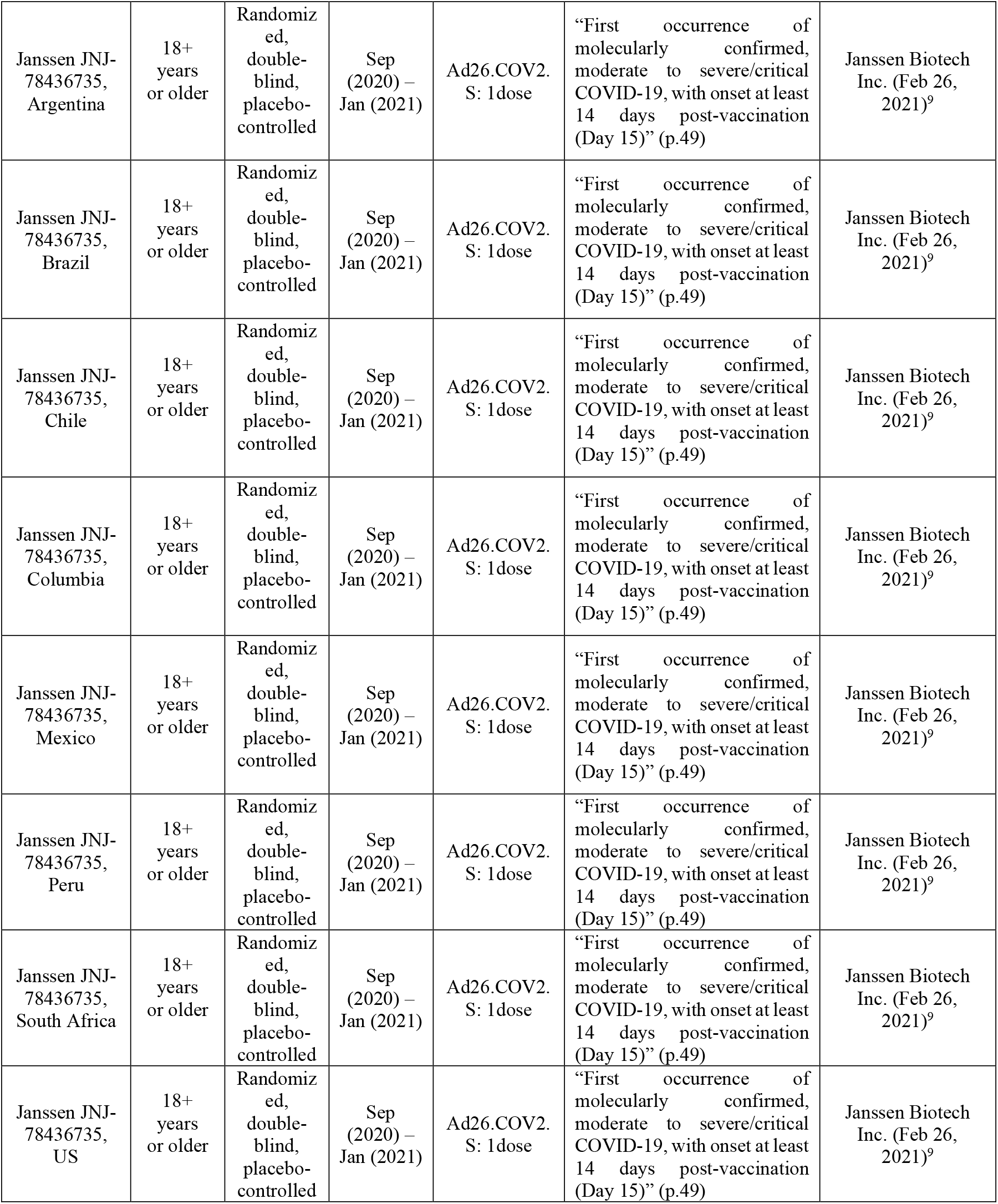
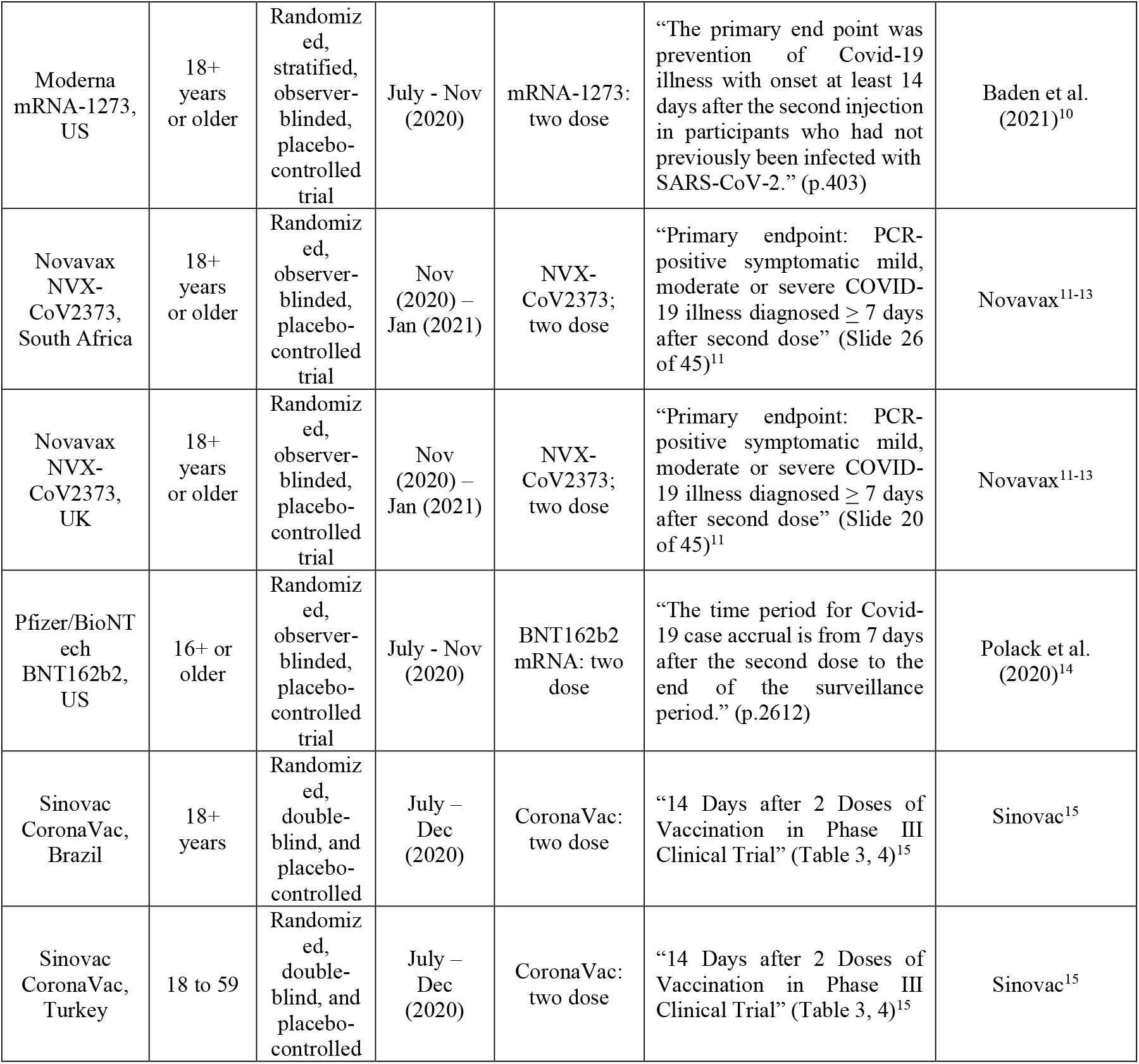
Primary data from Phase 3 Trials of SARS-CoV-2 vaccine candidates reporting efficacies.

| Vaccine Candidate | Population | Study type | Trial dates | Intervention | Primary endpoint for efficacy computation | Source |
| --- | --- | --- | --- | --- | --- | --- |
| AstraZeneca AZD1222, Brazil (SD/SD) | 18+ years | Randomized, blinded, controlled trials | April – Nov (2020) | ChAdOx1 nCoV-19: two dose | “The primary end point was prevention of Covid-19 illness with onset at least 14 days after the second injection in participants who had not previously been infected with SARS-CoV-2” (p.104) | Voysey et al (2021)<sup>7</sup> |
| AstraZeneca AZD1222-US, Chile and Peru | 18+ years | Randomized, double-blind, placebo-controlled | Aug (2020) – March (2021) | ChAdOx1 nCoV-19: two dose | “The primary endpoint, vaccine efficacy at preventing symptomatic COVID-19 was 76% (confidence interval (CI): 68% to 82%) occurring 15 days or more after receiving two doses given four weeks apart.” | AstraZeneca. AZD1222 US Phase III primary analysis confirms safety and efficacy (https://www.astrazeneca.com/media-centre/press-releases/2021/azd1222-us-phase-iii-primary-analysis-confirms-safety-and-efficacy.html). March 25, 2021. |
| AstraZeneca AZD1222, UK (LD/SD) | 18+ years | Randomized, blinded, controlled trials | April – Nov (2020) | ChAdOx1 nCoV-19: two dose | “The primary end point was prevention of Covid-19 illness with onset at least 14 days after the second injection in participants who had not previously been infected with SARS-CoV-2” (p.104) | Voysey et al (2021)<sup>7</sup> |
| AstraZeneca AZD1222, UK (SD/SD) | 18+ years | Randomized, blinded, controlled trials | April – Nov (2020) | ChAdOx1 nCoV-19: two dose | “The primary end point was prevention of Covid-19 illness with onset at least 14 days after the second injection in participants who had not previously been infected with SARS-CoV-2” (p.104) | Voysey et al (2021)<sup>7</sup> |
| Bharat Biotech, COVAXIN, India | 18-98 years | Randomized, double-blind, Placebo-controlled | Nov (2020) – Feb (2021) | BBV152: two dose | “The primary endpoint of Phase 3 clinical trial is based on the first occurrence of PCR-confirmed symptomatic (mild, moderate, or severe) COVID-19 with onset at least 14 days after the second study vaccination in serologically negative (to SARS-CoV-2) adult participants at baseline.” | https://www.bharatbiotech.com/images/press/covaxin-phase3-efficacy-results.pdf |
| Gamaleya rAd26/rAd5, Russia | 18+ years | Randomized, double-blind, placebo-controlled | Sept – Nov (2020) | rAd26/rAd5: two dose | “The primary outcome was the proportion of participants with PCR-confirmed COVID-19 from day 21 after receiving the first dose.” (p.671, 675) | Logunov et al (2021)<sup>8</sup> |
| Janssen JNJ-78436735, Argentina | 18+ years or older | Randomized, double-blind, placebo-controlled | Sep (2020) – Jan (2021) | Ad26.COV2. S: 1dose | “First occurrence of molecularly confirmed, moderate to severe/critical COVID-19, with onset at least 14 days post-vaccination (Day 15)” (p.49) | Janssen Biotech Inc. (Feb 26, 2021)<sup>9</sup> |
| Janssen JNJ-78436735, Brazil | 18+ years or older | Randomized, double-blind, placebo-controlled | Sep (2020) – Jan (2021) | Ad26.COV2. S: 1dose | “First occurrence of molecularly confirmed, moderate to severe/critical COVID-19, with onset at least 14 days post-vaccination (Day 15)” (p.49) | Janssen Biotech Inc. (Feb 26, 2021)<sup>9</sup> |
| Janssen JNJ-78436735, Chile | 18+ years or older | Randomized, double-blind, placebo-controlled | Sep (2020) – Jan (2021) | Ad26.COV2. S: 1dose | “First occurrence of molecularly confirmed, moderate to severe/critical COVID-19, with onset at least 14 days post-vaccination (Day 15)” (p.49) | Janssen Biotech Inc. (Feb 26, 2021)<sup>9</sup> |
| Janssen JNJ-78436735, Columbia | 18+ years or older | Randomized, double-blind, placebo-controlled | Sep (2020) – Jan (2021) | Ad26.COV2. S: 1dose | “First occurrence of molecularly confirmed, moderate to severe/critical COVID-19, with onset at least 14 days post-vaccination (Day 15)” (p.49) | Janssen Biotech Inc. (Feb 26, 2021)<sup>9</sup> |
| Janssen JNJ-78436735, Mexico | 18+ years or older | Randomized, double-blind, placebo-controlled | Sep (2020) – Jan (2021) | Ad26.COV2. S: 1dose | “First occurrence of molecularly confirmed, moderate to severe/critical COVID-19, with onset at least 14 days post-vaccination (Day 15)” (p.49) | Janssen Biotech Inc. (Feb 26, 2021)<sup>9</sup> |
| Janssen JNJ-78436735, Peru | 18+ years or older | Randomized, double-blind, placebo-controlled | Sep (2020) – Jan (2021) | Ad26.COV2. S: 1dose | “First occurrence of molecularly confirmed, moderate to severe/critical COVID-19, with onset at least 14 days post-vaccination (Day 15)” (p.49) | Janssen Biotech Inc. (Feb 26, 2021)<sup>9</sup> |
| Janssen JNJ-78436735, South Africa | 18+ years or older | Randomized, double-blind, placebo-controlled | Sep (2020) – Jan (2021) | Ad26.COV2. S: 1dose | “First occurrence of molecularly confirmed, moderate to severe/critical COVID-19, with onset at least 14 days post-vaccination (Day 15)” (p.49) | Janssen Biotech Inc. (Feb 26, 2021)<sup>9</sup> |
| Janssen JNJ-78436735, US | 18+ years or older | Randomized, double-blind, placebo-controlled | Sep (2020) – Jan (2021) | Ad26.COV2. S: 1dose | “First occurrence of molecularly confirmed, moderate to severe/critical COVID-19, with onset at least 14 days post-vaccination (Day 15)” (p.49) | Janssen Biotech Inc. (Feb 26, 2021)<sup>9</sup> |
| Moderna<br>mRNA-1273,<br>US | 18+<br>years<br>or older | Randomiz<br>ed,<br>stratified,<br>observer-<br>blinded,<br>placebo-controlled<br>trial | July - Nov<br>(2020) | mRNA-1273:<br>two dose | “The primary end point was prevention of Covid-19 illness with onset at least 14 days after the second injection in participants who had not previously been infected with SARS-CoV-2.” (p.403) | Baden et al.<br>(2021)<sup>10</sup> |
| Novavax<br>NVX-CoV2373,<br>South Africa | 18+<br>years<br>or older | Randomiz<br>ed,<br>observer-<br>blinded,<br>placebo-controlled<br>trial | Nov<br>(2020) –<br>Jan (2021) | NVX-CoV2373;<br>two dose | “Primary endpoint: PCR-positive symptomatic mild, moderate or severe COVID-19 illness diagnosed ≥ 7 days after second dose” (Slide 26 of 45)<sup>11</sup> | Novavax<sup>11-13</sup> |
| Novavax<br>NVX-CoV2373,<br>UK | 18+<br>years<br>or older | Randomiz<br>ed,<br>observer-<br>blinded,<br>placebo-controlled<br>trial | Nov<br>(2020) –<br>Jan (2021) | NVX-CoV2373;<br>two dose | “Primary endpoint: PCR-positive symptomatic mild, moderate or severe COVID-19 illness diagnosed ≥ 7 days after second dose” (Slide 20 of 45)<sup>11</sup> | Novavax<sup>11-13</sup> |
| Pfizer/BioNT<br>ech<br>BNT162b2,<br>US | 16+ or<br>older | Randomiz<br>ed,<br>observer-<br>blinded,<br>placebo-controlled<br>trial | July - Nov<br>(2020) | BNT162b2<br>mRNA: two<br>dose | “The time period for Covid-19 case accrual is from 7 days after the second dose to the end of the surveillance period.” (p.2612) | Polack et al.<br>(2020)<sup>14</sup> |
| Sinovac<br>CoronaVac,<br>Brazil | 18+<br>years | Randomiz<br>ed,<br>double-blind, and<br>placebo-controlled | July –<br>Dec<br>(2020) | CoronaVac:<br>two dose | “14 Days after 2 Doses of Vaccination in Phase III Clinical Trial” (Table 3, 4)<sup>15</sup> | Sinovac<sup>15</sup> |
| Sinovac<br>CoronaVac,<br>Turkey | 18 to 59 | Randomiz<br>ed,<br>double-blind, and<br>placebo-controlled | July –<br>Dec<br>(2020) | CoronaVac:<br>two dose | “14 Days after 2 Doses of Vaccination in Phase III Clinical Trial” (Table 3, 4)<sup>15</sup> | Sinovac<sup>15</sup> |

**eFigure 2:**
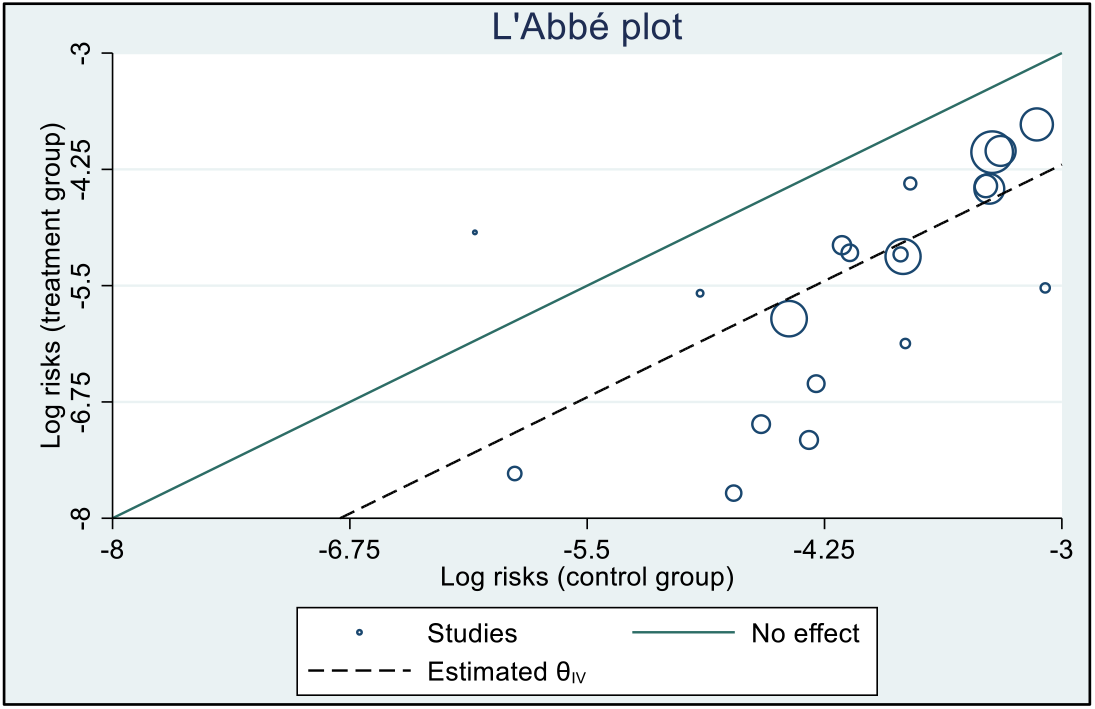
L’Abbe’ plot to explore heterogeneity.

**eFigure 3:**
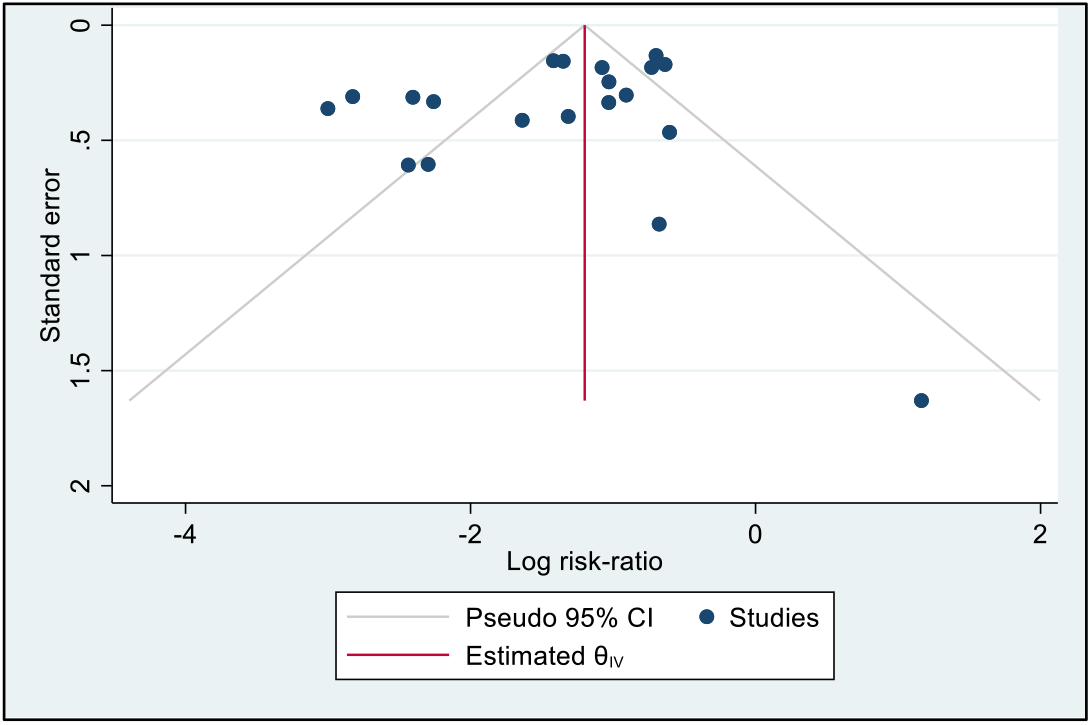
Funnel Plot.

**eTable 4:**
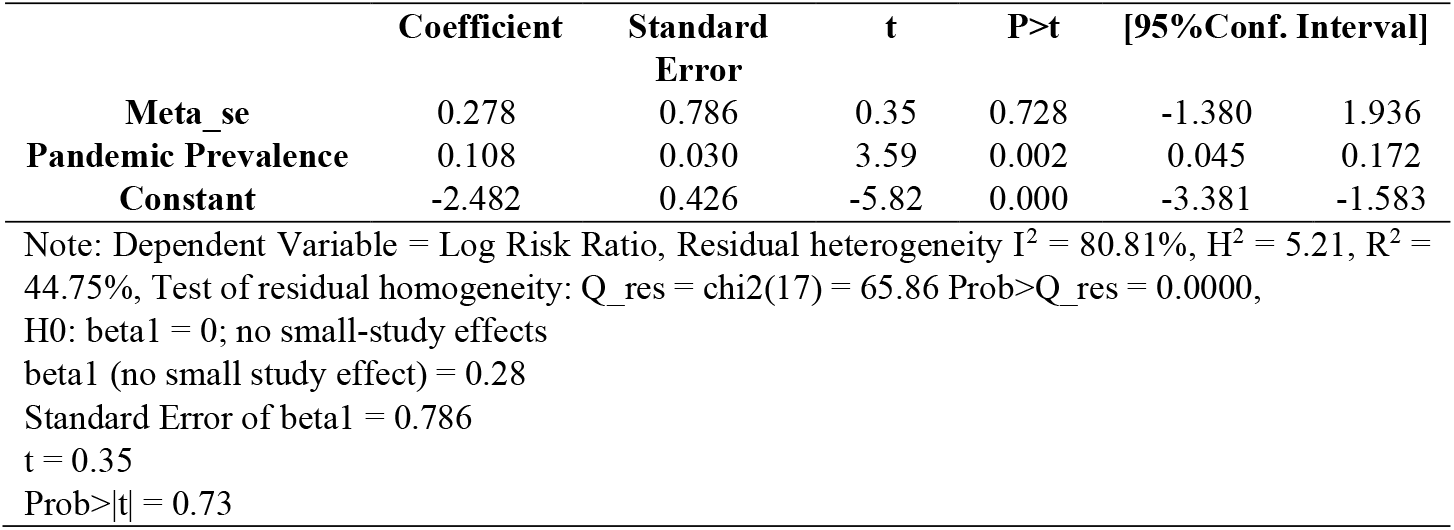
Results of Meta-regression –Egger Test.

**eTable 5:**
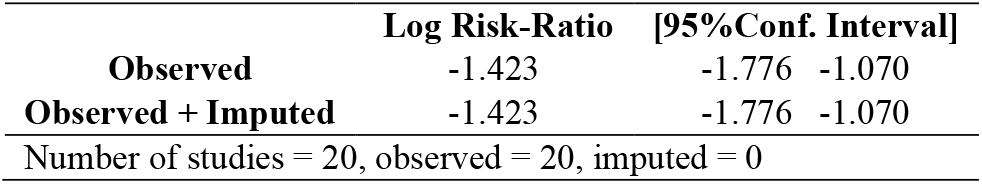
Non-parametric trim and fill test.

|  | Log Risk-Ratio | [95%Conf. Interval] |  |
| --- | --- | --- | --- |
| **Observed** | -1.423 | -1.776 | -1.070 |
| **Observed + Imputed** | -1.423 | -1.776 | -1.070 |

Number of studies = 20, observed = 20, imputed = 0

**eTable 6:** Results of Meta-regression including JanssenJNJ as a dummy variable.

|  | Coefficient | Standard<br>Error | t | P>t | [95%Conf. Interval] |  |
| --- | --- | --- | --- | --- | --- | --- |
| **Pandemic Prevalence** | 0.091 | 0.032 | 2.85 | 0.011 | 0.024 | 0.159 |
| **JanssenJNJ** | 0.319 | 0.323 | 0.99 | 0.337 | -0.362 | 1.10 |
| **Constant** | -2.369 | 0.296 | -7.98 | 0.000 | -2.995 | -1.742 |

Note: Dependent Variable = Log Risk Ratio, Residual heterogeneity $I^2 = 79.03\%$ , $H^2 = 4.77$ , $R^2 = 45.89\%$ , Test of residual homogeneity: $Q_{res} = \chi^2(17) = 67.71$ Prob> $Q_{res} = 0.0000$

**eTable 7:**
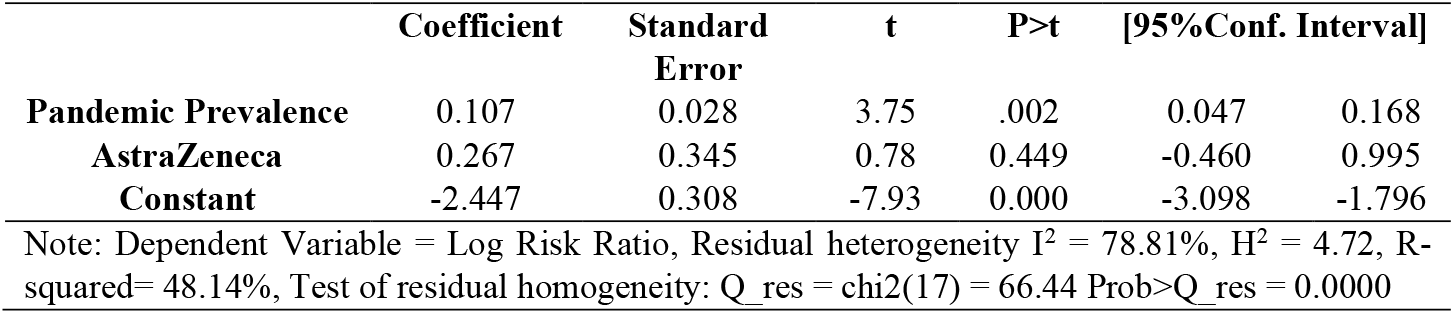
Results of Meta-regression including AstraZeneca as a dummy variable.

**eTable 8:** Results of Meta-regression – excluding three trials conducted in Brazil.

|  | Coefficient | Standard<br>Error | t | P>t | [95%Conf. Interval] |  |
| --- | --- | --- | --- | --- | --- | --- |
| **Pandemic Prevalence** | 0.137 | 0.041 | 3.29 | 0.005 | 0.048 | 0.227 |
| **Constant** | -2.54 | 0.349 | -7.27 | 0.000 | -3.289 | -1.798 |

Note: Dependent Variable = Log Risk Ratio, Residual heterogeneity $I^2 = 79.64\%$ , $H^2 = 4.91$ , $R^2 = 44.89\%$ , Test of residual homogeneity: $Q_{res} = \chi^2(15) = 55.2$ Prob> $Q_{res} = 0.0000$

**eTable 9:**
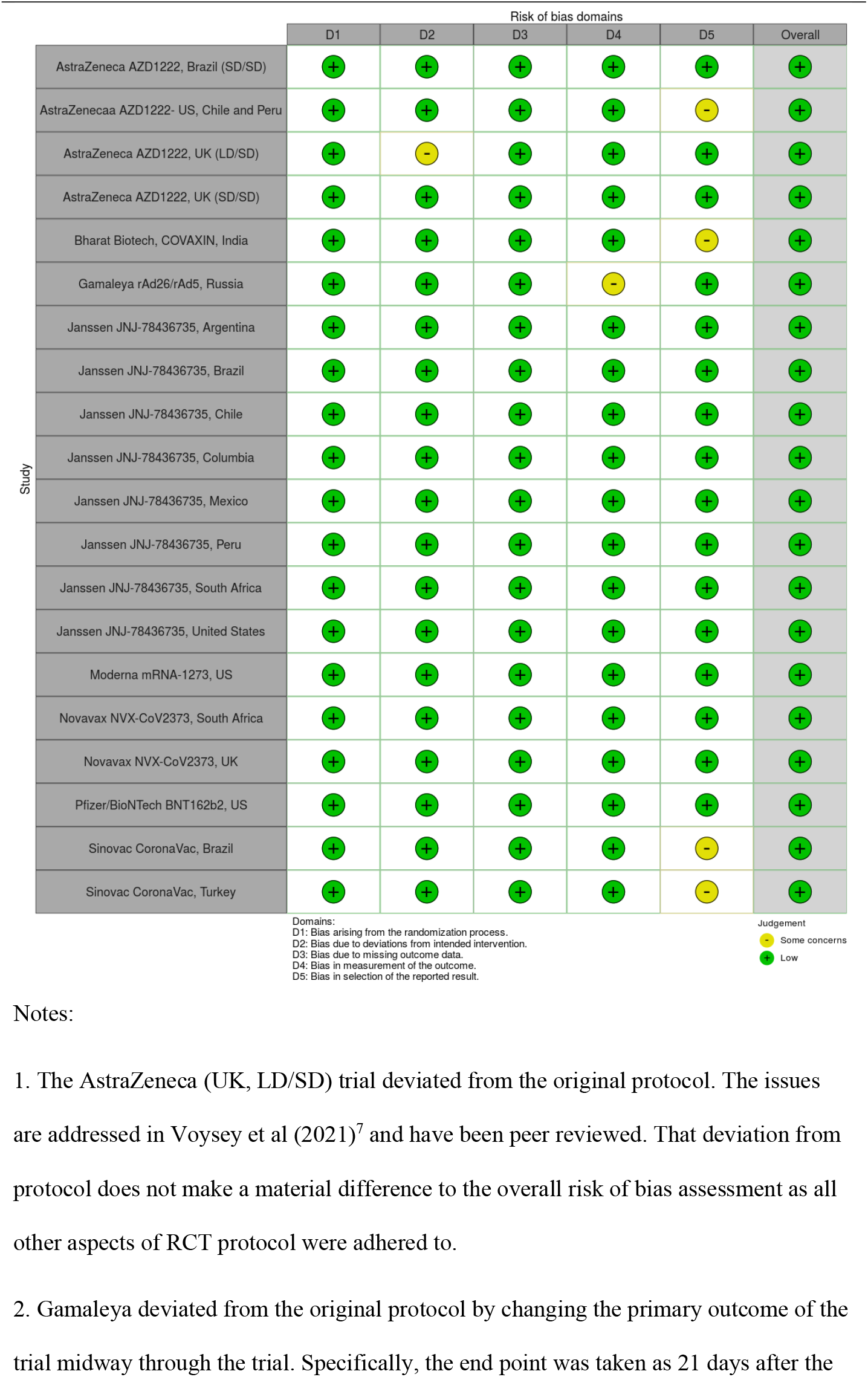
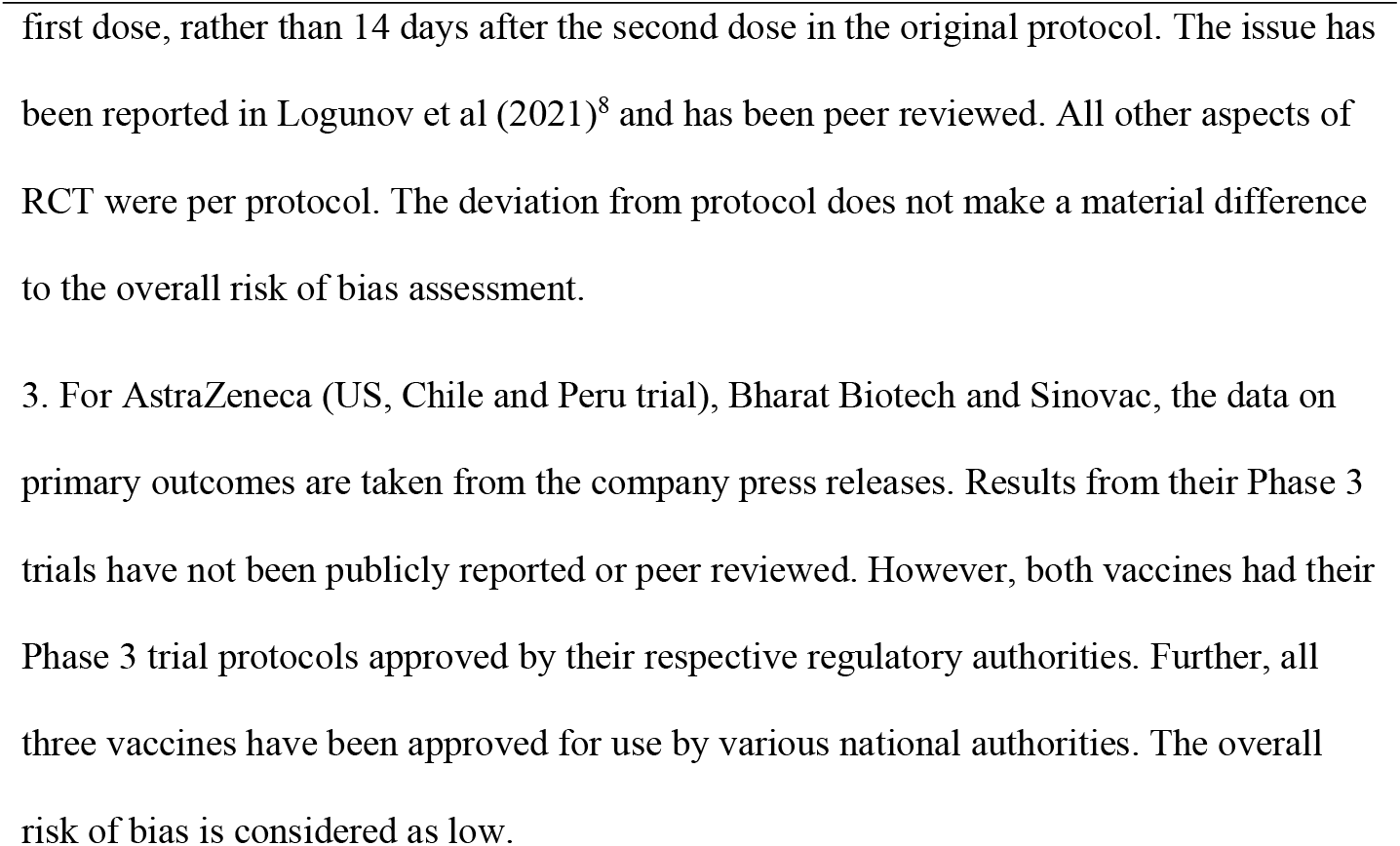
Risk of Bias Assessment.

**eTable 10:**
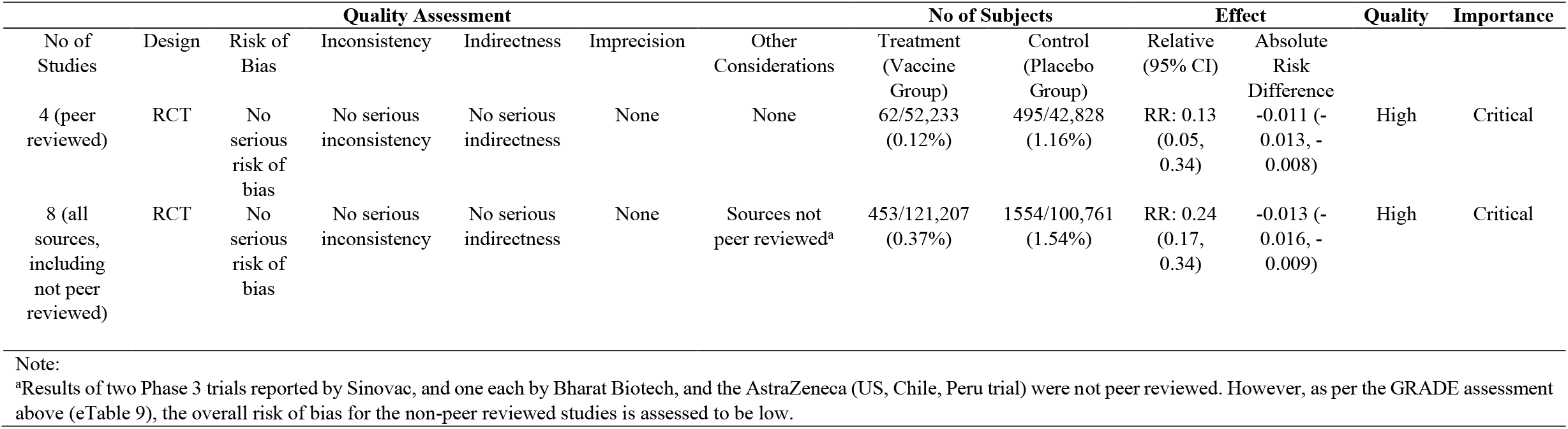
GRADE Assessment.

